# Development and Validation of a Parsimonious Financial Well-Being Index in the Health and Retirement Study

**DOI:** 10.64898/2026.10.18.26365842

**Authors:** Mina Habib, Elizabeth Rose Mayeda, Luke Barry, Onyebuchi A. Arah, Claudia L. Nau, Hua Zhou, Katrina L Kezios, Roch A. Nianogo

## Abstract

Financial well-being (FWB) encompasses objective and subjective financial status and may influence health. We developed and validated a parsimonious FWB index using data from 23,006 Health and Retirement Study (HRS) participants aged ≥50 years during 2006–2022. The FWB-5 Index was derived using correlated two-factor confirmatory factor analysis accounting for household clustering. Indicators included household wealth, household income-to-poverty ratio, monthly financial strain, satisfaction with one’s financial situation, and perceived financial mastery. We assessed convergent and discriminant validity using the Consumer Financial Protection Bureau Financial Well-Being (CFPB-FWB) Scale and Satisfaction with Life Scale, respectively. Associations with pre-baseline socioeconomic factors and subsequent depressive symptoms and health conditions were examined to assess construct and predictive validity. Model fit indices supported the two-factor structure (robust comparative fit index = 0.99; robust root mean square error of approximation = 0.05). Correlations were stronger with the CFPB-FWB Scale (Spearman ρ = 0.80) than with life satisfaction (ρ = 0.43). Associations with socioeconomic factors and subsequent health outcomes were in the hypothesized directions. These findings support the FWB-5 Index as a parsimonious measure for examining financial well-being and its associations with health across HRS waves, including those predating the CFPB-FWB Scale.

## Introduction

Financial well-being (FWB) has emerged as an important construct in health research because conventional measures of financial status, such as income and wealth, may not fully capture individuals’ financial circumstances and their implications for health. ^1–5^ Unlike these traditional measures, FWB is a holistic measure of financial status that encompasses both objective financial resources and subjective perceptions of financial security, control, and the ability to meet current and future financial obligations. ^1,2,4,5^ Individuals with similar levels of income or wealth may experience substantially different levels of financial strain, perceived security, and capacity to manage unexpected expenses, potentially influencing health-related behaviors, psychological well-being, and access to resources. ^1,2,4,5^ These dimensions may be particularly relevant during midlife and older adulthood, when retirement, healthcare expenditures, and caregiving responsibilities can substantially alter financial circumstances. Despite its potential importance for understanding health and aging, longitudinal research on FWB has been constrained by the limited availability of standardized measures in established aging cohorts. In 2015, the U.S. Consumer Financial Protection Bureau (CFPB) advanced the conceptualization and standardized measurement of FWB through the development of the CFPB Financial Well-Being Scale (CFPB-FWB). ^2,5^ However, the relatively recent introduction of this measure limits opportunities to investigate long-term associations between FWB and health outcomes using historical data from longitudinal aging studies.

Repeated measures of FWB have also been recommended for research examining cumulative financial exposures, changes over time, social mobility, and the timing of financial circumstances across the life course, ^1^ underscoring the importance of longitudinal assessment. Large-scale cohort studies have begun incorporating measures of the construct. ^6,7^ In particular, in 2020 the HRS, a large ongoing longitudinal study of U.S. adults aged 51 years and older and their spouses, included the 10 items used to calculate the CFPB-FWB Scale in its Leave-Behind Psychosocial and Lifestyle Questionnaire. ^8^ Although this represents an important advancement, the recent introduction of these measures limits longitudinal data availability for these well-established cohorts.

Previous research has strived to address this limitation by constructing alternative measures of FWB using existing HRS data. ^4^ Kezios et al. (2026) developed an eight-item FWB index and calibrated it against the CFPB-FWB Scale.^4^ This work provides an important foundation for assessing FWB longitudinally in the HRS. This study builds on this important work by addressing additional methodological considerations. Currently the index relies on dichotomized indicators combined using an unweighted sum, which can reduce information contained in the original measures and assumes that each indicator contributes equally to FWB. In addition, conceptually related indicators may capture overlapping aspects of financial circumstances, potentially giving greater weight to domains represented by multiple items. Finally, the index is constructed from 8-items, but the authors suggest the possibility that a reduced index may be adequate while allowing for potential harmonization with other population-based studies that include these items. These gaps suggest an opportunity to develop a more parsimonious, model-based measure of FWB using indicators that are consistently available across HRS waves.

Accordingly, this study aims to develop and validate a parsimonious financial well-being index using five consistently assessed HRS indicators. We use a confirmatory factor analysis (CFA) framework to estimate an underlying FWB construct based on the relationships among the indicators. We evaluate the convergent validity of a 5-item FWB Index (“FWB-5” or “FWB-5 Index”) against the CFPB-FWB Scale, assess discriminant validity using the Satisfaction with Life Scale, judge construct validity through associations with theoretically related socioeconomic and psychosocial measures, and examine predictive validity in relation to subsequent health outcomes. The resulting FWB-5 Index will provide a parsimonious measure that can support longitudinal research on financial well-being, health, and aging.

## Methods

### Study Population and Data Sources

We used data from the Health and Retirement Study. First, we used data from the biennial HRS core interviews conducted from 2006 through 2022. ^7,9^ Second, we used data from the Psychosocial and Lifestyle Questionnaire, also referred to as the Leave-Behind Questionnaire (LBQ), administered from 2006 through 2022 to alternating subsets of HRS participants. ^7,10^ Although components of the proposed index were collected starting in the 2004 LBQ, we used 2006 as the starting wave because 2004 was a pilot year that preceded the formal implementation of the LBQ and its alternating random half-sample design. Beginning in 2006, the LBQ was integrated into the enhanced face-to-face interview and administered to alternating random 50% subsamples across waves. Third, we used the Cross-Wave Childhood Health and Family Aggregated Data file, which combines participant-reported information on childhood family circumstances and health collected across HRS waves from 1992 through 2016. ^11^ The HRS is sponsored by the National Institute on Aging (NIA U01AG009740) and conducted by the University of Michigan. Additional details on the HRS study design and data collection procedures have been published elsewhere. ^7,12^

As shown in **Figure 1**, three analytic samples were constructed among participants aged 50 years and older who were eligible for and completed the LBQ between 2006 and 2022. These samples were defined according to data availability and the specific objectives of the measurement development and validation analyses.

**Figure 1.**
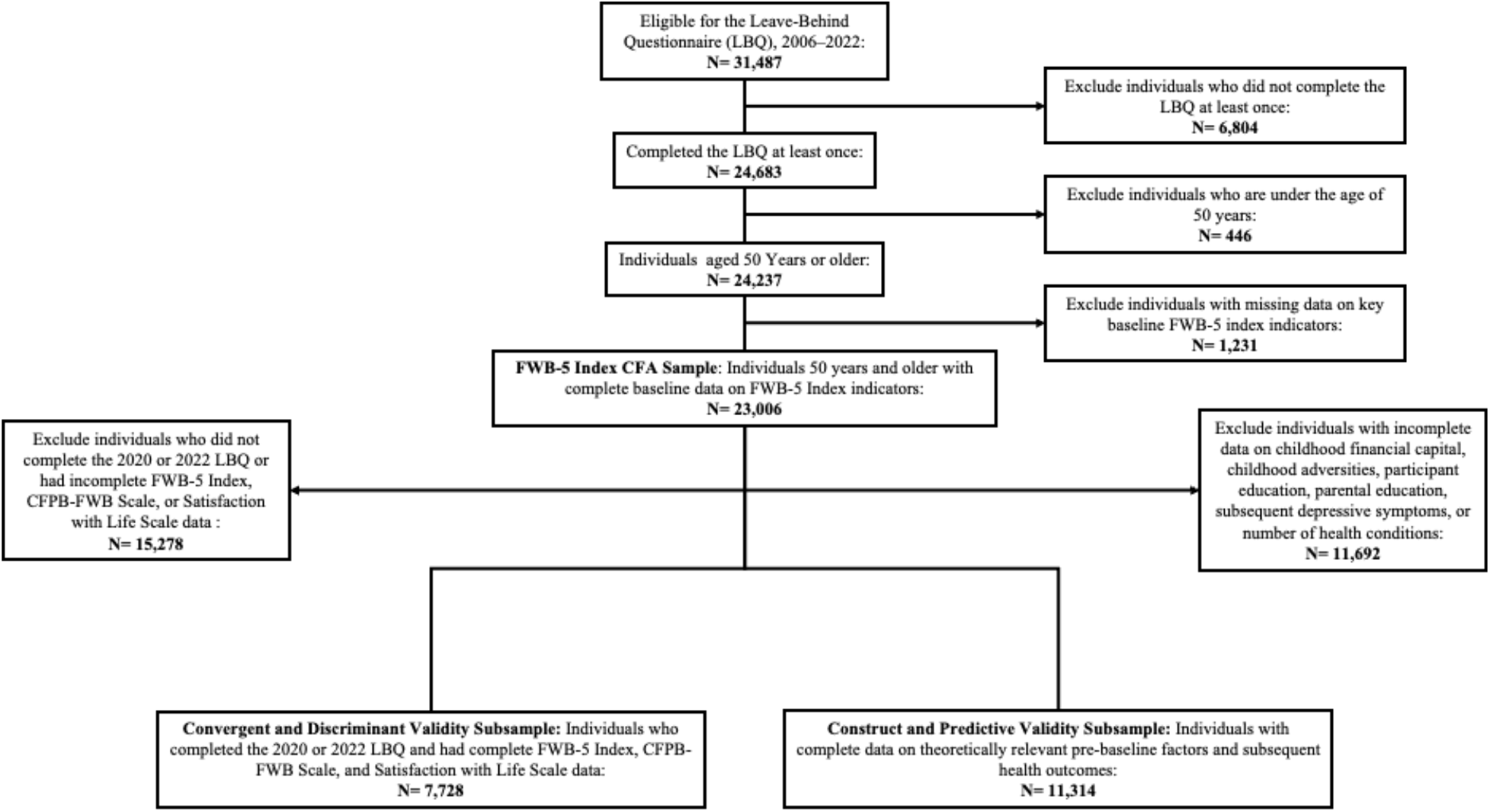
Flowchart of Analytical Samples, Health and Retirement Study, 2006–2022.

The first sample, the ‘FWB-5 Index Development and Validation Sample’, included 23,006 participants from 16,280 households who completed the LBQ at least once between 2006 and 2022 and had complete data on all five indicators used to construct the FWB-5. The sample included an average of 1.41 participants per household. Each participant’s first completed LBQ was designated as their analytic baseline, with subsequent observations aligned according to time since baseline. This sample was used to develop and validate the FWB-5 Index through confirmatory factor analysis. The resulting measurement model was subsequently applied to repeated LBQ observations to generate longitudinal FWB-5 scores, as described in the Statistical Analysis section.

The second sample, the ‘Convergent and Discriminant Validity Subsample’, included 7,728 participants from the first sample who completed the LBQ in 2020 or 2022 and had sufficient data to calculate the CFPB-FWB and Satisfaction with Life Scale scores. These participants were included to evaluate convergent validity against the CFPB-FWB Scale and discriminant validity against the Satisfaction with Life Scale. For these analyses, FWB-5 Index scores and the corresponding validation measures were calculated using data from the same 2020 or 2022 interview, rather than the participant’s initial LBQ baseline.

The third sample, the ‘Construct and Predictive Validity Subsample’, included 11,314 participants from the first sample with complete data on theoretically relevant pre-baseline characteristics and subsequent health outcomes. This sample was used to examine associations between the FWB-5 Index and theoretically related characteristics (construct validity), as well as subsequent health outcomes (predictive validity).

### Variables and Measures

#### Consumer Financial Protection Bureau Financial Well-Being Scale

The CFPB-FWB Scale is a ten-item measure of perceived financial well-being.^5^ The first six items assess how well statements describe the participant or their financial situation, with response options ranging from “Completely” to “Not at all”: 1) *I could handle a major unexpected expense*, 2) *I am securing my financial future*, 3) *Because of my money situation, I feel like I will never have the things I want in life*, 4) *I can enjoy life because of the way I’m managing my money*, 5) *I am just getting by financially*, and 6) *I am concerned that the money I have or will save won’t last*. The remaining 4 items assess how often statements about the participant’s financial situation apply, with response options ranging from “Always” to “Never”: 7) *Giving a gift for a wedding, birthday or other occasion would put a strain on my finances for the month*, 8) *I have money left over at the end of the month*, 9) *I am behind with my finances*, and 10) *My finances control my life*. Item responses were scored from zero to four, with negatively worded items reverse-coded such that higher values consistently represented greater financial well-being. Items three, five, six, seven, nine, and ten were reverse-coded in accordance with CFPB scoring procedures. Item scores were summed to obtain a raw total score, which was then converted to a CFPB-FWB score using the CFPB scoring table specific to age group and survey administration mode. Administration mode was determined using the LBQ indicator, which distinguishes self-administered questionnaires from interviewer-administered questionnaires. The resulting score ranges from zero to 100, with higher scores indicating greater financial well-being. Scores for participants missing any of the ten items were discarded. The ten CFPB-FWB items were included in the 2020 and 2022 HRS LBQs.^13^ Supplementary **Table S2** presents the CFPB-FWB Scale items, response options, and CFPB-FWB Scale score. The CFPB-FWB Scale score was standardized to facilitate comparison with the FWB-5 Index.

#### Objective and subjective indicators used to construct the FWB-5 Index

Consistent with conceptualizations of FWB as encompassing objective and subjective domains,^14^ we selected five indicators of objective and subjective financial status. The objective financial status domain included total household wealth, adjusted for inflation using the Consumer Price Index and expressed in 2022 dollars, and the household income-to-poverty ratio. Both measures were obtained from the HRS core interview and represent accumulated financial resources and household income relative to the poverty threshold, respectively. Both indicators were winsorized at the 1st and 99th percentiles to reduce the influence of extreme values.

The subjective financial status domain included three items from the LBQ: monthly financial strain, perceived financial mastery, and satisfaction with one’s present financial situation. Monthly financial strain was assessed by asking participants, “How difficult is it for you or your family to meet monthly payments on your or your family’s bills?” Responses ranged from 1 (“Not at all difficult”) to 5 (“Completely difficult”) and were reverse-coded so that higher values indicated less financial strain. Perceived financial mastery was assessed by asking participants to rate their control over their financial situation on a scale from 0 (“No control at all”) to 10 (“Very much control”), with higher scores indicating greater perceived control. Satisfaction with one’s present financial situation was assessed on a scale from 1 (“Not at all satisfied”) to 5 (“Completely satisfied”), with higher scores indicating greater satisfaction. All subjective indicators were assessed at the individual level and could therefore differ among members of the same household.

Indicator selection also prioritized consistent measurement across study waves. Although Kezios et al. included an item assessing distress related to ongoing financial strain from the general ongoing chronic stress scale, we excluded this item because it was not assessed in 2008. We also excluded “taking less medication because of cost,” given its limited convergence with the CFPB-FWB items in prior work and potential overlap with the selected indicators of financial strain.

#### Additional measures and covariates

To characterize the analytic sample, we included age (continuous, years), sex (male or female), marital status (binary, 0 = Not married, 1 = Married), and race and ethnicity (Hispanic, non-Hispanic Black, non-Hispanic White, or other race or ethnicity). For construct and predictive validity analyses, we additionally considered theoretically relevant measures of early-life socioeconomic conditions, education, mental health, and physical health.

Childhood financial capital index, developed by Vable et al. using HRS core and LBQ data, summarizes childhood financial resources and financial stability.^15^ This measure is akin to a child FWB index and higher values on the childhood financial capital index indicated greater childhood financial well-being across both objective and subjective dimensions. The childhood financial capital index was standardized to our baseline sample distribution. We additionally included participant educational attainment (continuous, years), parental educational attainment (continuous, years), and the number of childhood adversities experienced before age 18. Childhood adversity was operationalized as a count ranging from 0 to 4 based on whether participants reported having: 1) *repeated a year of school*, 2) *been in trouble with the police*, 3) *lived with a parent whose alcohol or drug use caused problems in the family*, or 4) *been physically abused by either parent*.

Depressive symptoms were assessed using the shortened 8-item Center for Epidemiologic Studies Depression (CES-D) measure used in the HRS, with higher scores indicating a greater number of depressive symptoms (Range 0 to 8). Physical health burden was measured as a continuous count of 8 health conditions ever reported: hypertension or high blood pressure; diabetes or high blood sugar; cancer or a malignant tumor other than skin cancer; chronic lung disease other than asthma (e.g. chronic bronchitis or emphysema); heart attack, coronary heart disease, angina, congestive heart failure, or other heart problems; stroke or transient ischemic attack; emotional, nervous, or psychiatric problems; and arthritis or rheumatism (Range 0 to 8).

Finally, we assessed discriminant validity using Diener’s Satisfaction with Life Scale, a well-established measure of global life satisfaction included in the LBQ.^13^ Respondents rated their agreement with five statements, with responses ranging from “Strongly disagree” to “Strongly agree”: “In most ways my life is close to ideal,” “The conditions of my life are excellent,” “I am satisfied with my life,” “So far, I have gotten the important things I want in life,” and “If I could live my life again, I would change almost nothing.” Scores were calculated by averaging responses across the five items, with higher scores indicating greater life satisfaction.^13^

### Statistical Analysis

#### Descriptive statistics

We summarized the baseline characteristics of the primary FWB-5 Index CFA analytic sample in **Table 1**. For the convergent and discriminant validity and construct and predictive validity samples, we additionally summarized subsample-specific characteristics and relevant covariates in Supplementary **Tables S2** and **S3**.

**Table 1.** Baseline Characteristics of the FWB-5 Index Confirmatory Factor Analysis Sample, Health and Retirement Study, 2006–2022.

| Characteristic | N = 23,006<sup>1</sup> |
| --- | --- |
| Age (Years) | 65 (10) |
| Male | 9,735 (42%) |
| Race/Ethnicity |  |
| Hispanic | 2,826 (12%) |
| NH Black | 4,029 (18%) |
| NH White | 15,301 (67%) |
| Other | 819 (3.6%) |
| Unknown | 31 |
| Participant Years of Education | 12.8 (3.1) |
| Unknown | 85 |
| Married | 13,999 (61%) |
| Unknown | 9 |
| Mother's Education (Years) | 10.0 (3.7) |
| Unknown | 2,574 |
| Father's Education (Years) | 9.7 (4.0) |
| Unknown | 4,138 |
| Childhood Financial Capital Index Z-Score | 0.00 (1.00) |
| Unknown | 7,385 |
| Number of Childhood Adversities | 0.36 (0.67) |
| Std. FWB-5 Index | 0.00 (1.00) |
| Std. Subjective Financial Status Factor Score | 0.00 (1.00) |
| Financial Mastery Index | 7.19 (2.69) |
| End-of-Month Financial Strain |  |
| Not at all difficult | 8,322 (36%) |
| Not very difficult | 6,729 (29%) |
| Somewhat difficult | 5,729 (25%) |
| Very difficult | 1,621 (7.0%) |
| Completely difficult | 605 (2.6%) |
| Satisfaction with Present Financial Situation |  |
| Not at all satisfied | 2,180 (9.5%) |
| Not very satisfied | 3,740 (16%) |
| Somewhat satisfied | 8,091 (35%) |
| Very satisfied | 5,734 (25%) |
| Completely satisfied | 3,261 (14%) |
| Std. Objective Financial Status Factor Score | 0.00 (1.00) |
| Total Household Wealth (2022 U.S. Dollars) | 559,315 (932,509) |
| Household Income-to-Poverty Ratio | 4.8 (4.8) |
| Number of Conditions Ever Reported | 1.92 (1.45) |
| CES-D/Depression Symptoms | 1.50 (2.01) |
| Unknown | 485 |

<sup>1</sup> Mean (SD); n (%)

#### Confirmatory Factor Analysis Approach to Calculating the FWB-5 Index

The FWB-5 Index was developed using a correlated two-factor confirmatory factor analysis (CFA), consistent with conceptualizations of financial well-being as encompassing objective and subjective domains. ^2,4,14,15^ Prior psychometric research on the Multidimensional Subjective Financial Well-being Scale (MSFWBS), which used bifactor exploratory structural equation modeling (ESEM), further informed our consideration of the construct’s multidimensionality. CFA is commonly used in instrument development and validation when relationships between observed indicators and underlying latent factors are specified a priori.^16–20^ The objective financial status domain was represented by baseline total household wealth and household income-to-poverty ratio, both winsorized and standardized. Objective indicators entered the model as continuous numeric variables. The subjective financial status domain was represented by monthly financial strain, perceived financial mastery, and satisfaction with one’s present financial situation. Monthly financial strain was reverse-coded so that higher values indicated better financial well-being. Subjective indicators were entered into the model as continuous ordered variables.

Using baseline observations from the primary FWB-5 Index confirmatory factor analysis sample to avoid dependence from repeated measures, we randomly divided households into approximately equal groups, with all household members retained together, into equal training and test samples. We specified a correlated 2-factor CFA in which each observed indicator loaded onto its hypothesized latent domain. Models were estimated using maximum likelihood with robust standard errors (MLR) to account for potential departures from multivariate normality. We additionally accounted for clustering at the household level to accommodate potential within-household correlations, particularly because income and wealth were measured at the household level. Model fit was evaluated using standardized factor loadings and global fit indices, including the robust comparative fit index (CFI) and robust root mean square error of approximation (RMSEA). ^16–20^ Standardized factor loadings reflect the strength of the relationship between each observed indicator and its corresponding latent factor, with larger absolute values indicating stronger associations. The CFI assesses model fit relative to a null model, with values greater than 0.90 generally considered indicative of acceptable fit. RMSEA assesses approximate model fit while accounting for model complexity, with lower values indicating better fit.

We first evaluated the prespecified correlated 2-factor model in the test sample by examining factor loadings, CFI, RMSEA, and modification indices in the holdout sample.^18,20^ The model demonstrated good fit in the test sample (CFI=0.99; RMSEA= 0.05) and was subsequently refit using the full data. Training-validation factor loadings and fit statistics were similar. As a sensitivity analysis, we also fit a 1-factor model, which demonstrated substantially poorer fit (CFI=0.82; RMSEA=0.21) and was inconsistent with the hypothesized multidimensional structure of FWB. Consequently, we retained the evaluation of the index solely on the two-factor model. Full model-fit statistics and factor loadings for both models are presented in supplementary **Table S1**. Additionally, supplementary **Figure S1** presents the correlated 2-factor CFA path diagram for the holdout validation sample.^21^

Consistent with the approach used by Vable et. al. to construct the childhood financial capital index,^15^ the resulting objective and subjective factor domain scores were standardized, summed, and standardized once more to the baseline sample distribution. The FWB-5 Index is therefore a continuous standardized index, with higher scores indicating greater financial well-being. Supplementary **Figure S2** illustrates the steps used to construct and operationalize the FWB-5 Index.

#### Assessing Convergent and Discriminant Validity

Within the convergent and discriminant validity sample, we assessed the correlation between sample-standardized FWB-5 Index and CFPB-FWB Scale scores using Spearman rank correlation coefficients.^4,20,22^ This approach was consistent with the validation methodology used by Kezios et al. (2026).^4,22^ Similarly, we considered a Spearman correlation coefficient greater than 0.50 between the FWB-5 Index and CFPB-FWB Scale as evidence of convergent validity.

Although our primary aim was to assess convergence between the overall FWB-5 Index and CFPB-FWB Scale scores, we additionally conducted exploratory analyses to characterize correspondence between their underlying components. Specifically, we estimated Spearman correlations among the FWB-5 total score, its objective and subjective domain factor scores, the overall CFPB-FWB Scale score, and individual CFPB-FWB items. Negatively worded CFPB-FWB items were reverse coded such that higher responses indicated increased financial well-being. These analyses were used to identify which components of the CFPB-FWB Scale were most strongly represented by the FWB-5 Index. **Figure 3** presents the resulting correlation matrix.

Finally, we assessed discriminant validity, the extent to which a measure is distinct from measures of other constructs,^20^ using the Satisfaction with Life Scale. A weaker Spearman correlation with this scale than with the CFPB-FWB Scale was considered evidence consistent with discriminant validity.

#### Assessing Construct and Predictive Validity

Given that this is one of the first operationalizations of adult financial well-being in the HRS, we evaluated both the construct validity and the predictive validity of the FWB-5 Index.^4,15,23^ Construct validity refers to the extent to which the index correlates with theoretically relevant constructs. ^15,23^ On the basis of prior literature, we hypothesized that childhood financial capital index,^15,24^ the number of childhood adversities,^25^ participants’ years of education,^26^ and parental education,^27,28^ all of which occurred before the analytical baseline, would be associated with baseline standardized FWB-5 scores. Consistent with the approach used in developing the childhood financial capital index, we fitted separate unadjusted linear regression models with baseline standardized FWB-5 scores as the outcome and each hypothesized correlate entered individually as the predictor. ^15^ Beta coefficients were used to assess these associations. We also evaluated the predictive validity of the FWB-5 Index, which refers to the ability of a scale to predict future outcomes. ^15,23^ We hypothesized that depressive symptoms, measured using the validated CES-D scale, and the number of health conditions ever reported at the subsequent biennial study visit would be predicted by baseline standardized FWB-5 Index score. ^1,29,30^ To test this, we again fitted simple linear regression models in which each future outcome was regressed on baseline standardized FWB-5 Index score. Full regression results are presented in **Table 2**.

**Table 2.** Linear Regression Models Evaluating Associations Between the FWB Index and Theoretical Correlates in the Construct and Predictive Validity Sample, Health and Retirement Study, 2006–2022.

| Variable | Estimate | 95% CI | P-Value |
| --- | --- | --- | --- |
| **Panel A. Construct validity: Outcome = baseline standardized FWB-5 score** |  |  |  |
| Childhood Financial Capital Index Z-Score | 0.15 | 0.13, 0.17 | <0.001 |
| Number of Childhood Adversities | -0.17 | -0.2, -0.14 | <0.001 |
| Participant Years of Education | 0.11 | 0.1, 0.11 | <0.001 |
| Mother's Education | 0.05 | 0.05, 0.06 | <0.001 |
| Father's Education | 0.05 | 0.04, 0.05 | <0.001 |
| **Panel B. Predictive validity: Predictor = baseline standardized FWB-5 score** |  |  |  |
| Number of Conditions Ever Reported at Follow-up | -0.26 | -0.29, -0.24 | <0.001 |
| CES-D/Depression Score at Follow-up | -0.47 | -0.51, -0.44 | <0.001 |

#### Software

All statistical analyses were performed in R, version 4.5.2 (R Foundation for Statistical Computing, Vienna, Austria).^31^ Confirmatory factor analyses were conducted using the *lavaan* package.^32^

## Results

### Baseline Sample Characteristics

After applying the initial inclusion and exclusion criteria, the analytic sample included 23,006 participants who completed the LBQ at least once between 2006 and 2022 and had complete analytic baseline data for all FWB-5 Index financial indicators (**Figure 1**; **Table 1**). At baseline, the mean age was 65 years (SD = 10), 58% of participants were female, and 67% were non-Hispanic White. These characteristics were generally consistent across the two subsamples used for the convergent, construct, and predictive validity analyses, although mean age was slightly higher in subsamples using data collected after baseline (**Tables S2** and **S3**). Participants had a mean of 12.8 years of education (SD = 3.1), 61% were married, and their parents had approximately 10 years of education, on average. Participants reported fewer than one childhood adversity on average (mean = 0.36, SD = 0.67), fewer than 2 of 8 depressive symptoms (mean = 1.50, SD = 2.01), and fewer than 2 of 8 health conditions (mean = 1.92, SD = 1.45).

At baseline, the mean financial mastery score was 7.19 (SD = 2.69) out of 10, and 65% of participants reported that meeting monthly bills was either “not very difficult” or “not at all difficult.” 39% were “very satisfied” or “completely satisfied” with their present financial situation. For the objective financial indicators, mean winsorized total household wealth (in 2022 dollars) was $559,315 (SD = $932,509), and the mean winsorized household income-to-poverty ratio was 4.8 (SD = 4.8).

### Convergent and Discriminant Validity

Within the convergent and discriminant validity sample, the FWB-5 Index was strongly correlated with the CFPB-FWB Scale (Spearman ρ = 0.80, 95% CI: 0.79 to 0.81). The distributions of the standardized FWB-5 Index and CFPB-FWB scores shown in **Figure 2** provide a visual representation of this association. Furthermore, using a correlation threshold of 0.50, the FWB-5 Index was correlated with 8 of the 10 individual CFPB-FWB items (**Figure 3**). The remaining two items also showed moderate positive correlations with the FWB-5 Index: “Just getting by financially” (ρ = 0.46) and “Finances control my life” (ρ = 0.47).

**Figure 2.**
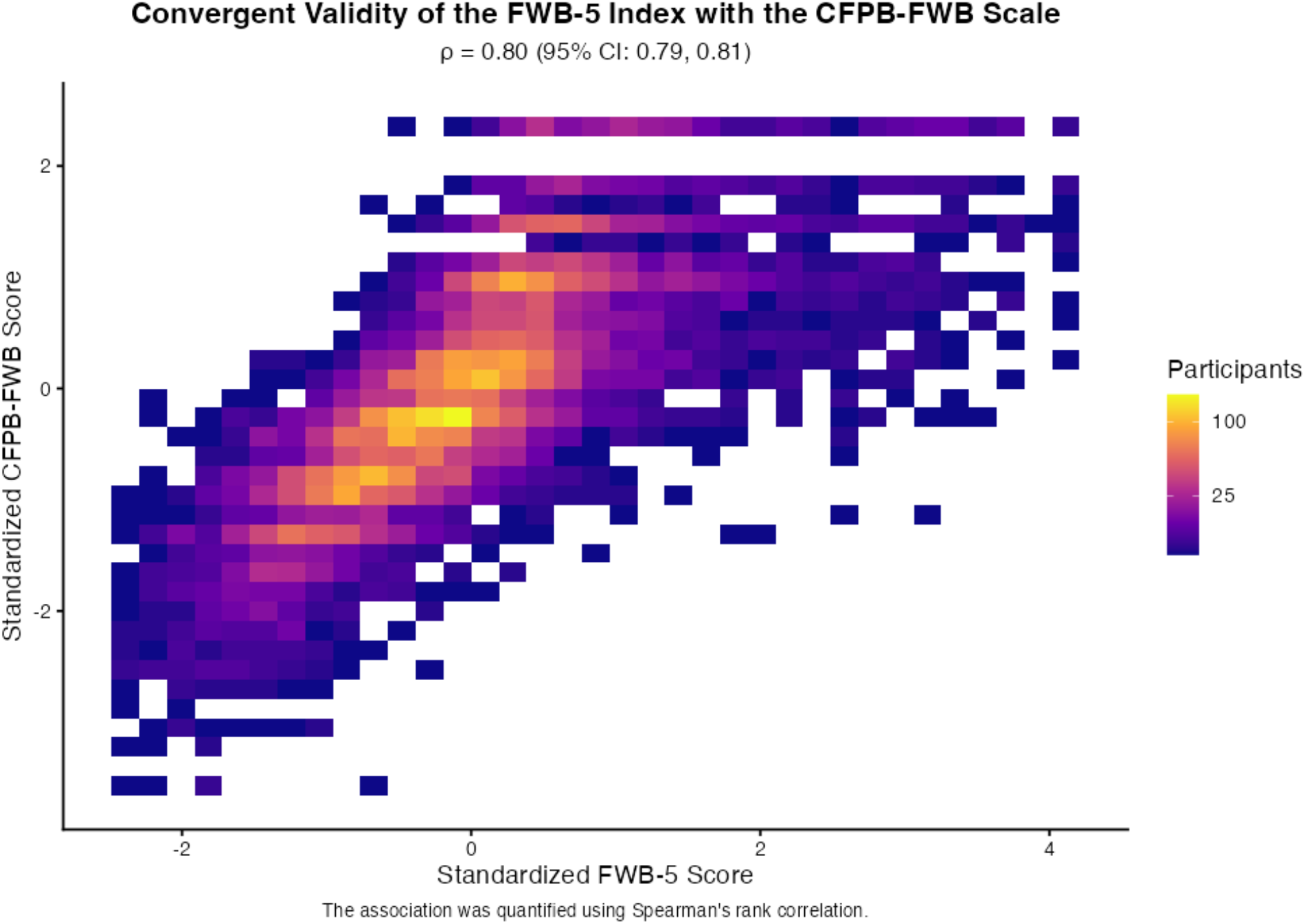
Correlation Between the CFPB-FWB Scale and the FWB-5 Index Score in the Convergent and Discriminant Validity Sample, Health and Retirement Study, 2020–2022.

**Figure 3.**
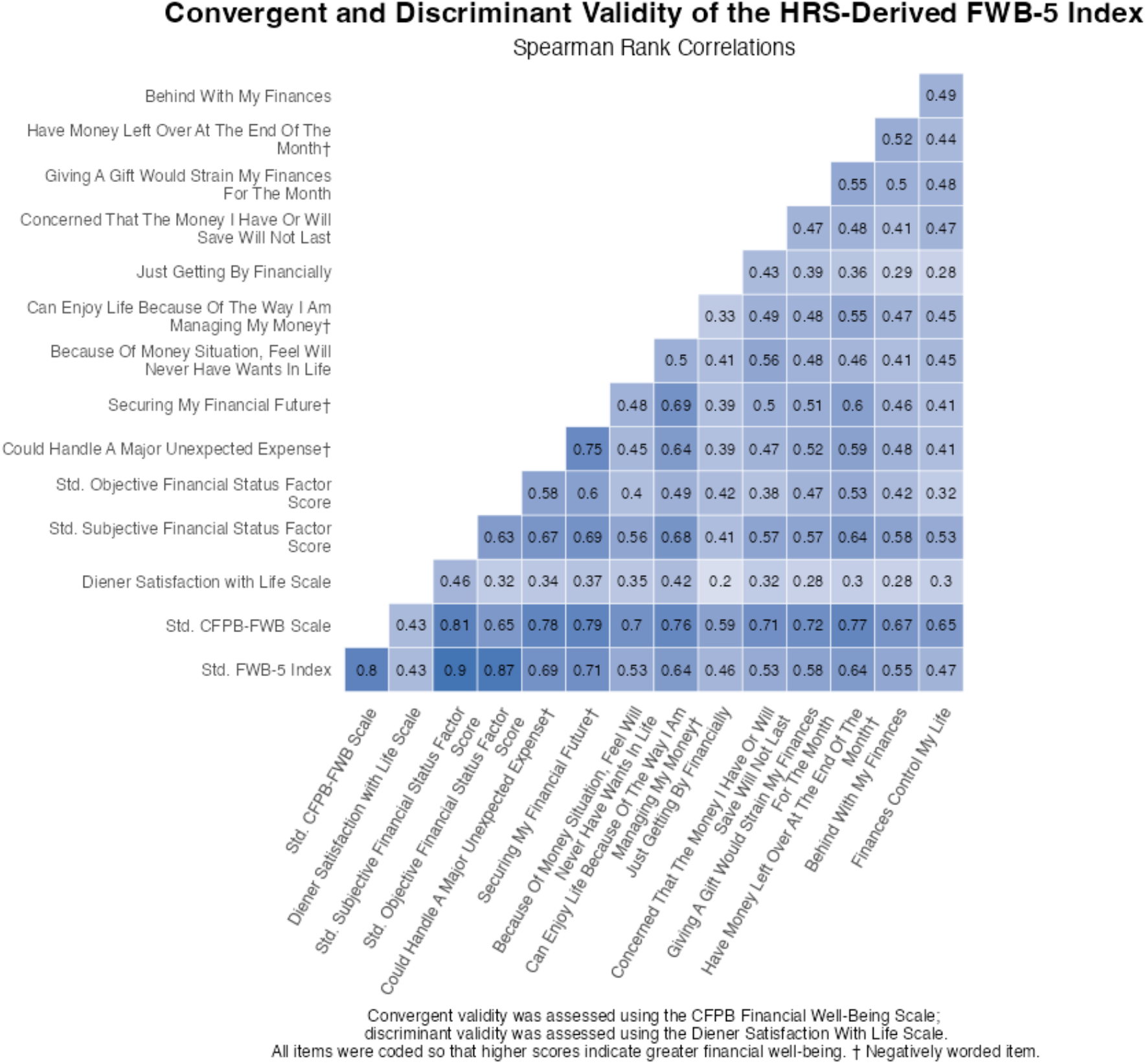
Correlation Matrix of the FWB-5 Index, CFPB-FWB Scale, and Component Measures in the Convergent and Discriminant Validity Sample, Health and Retirement Study, 2020–2022.

Both the FWB-5 Index and the CFPB-FWB Scale were more weakly correlated with the Satisfaction with Life Scale (Spearman ρ = 0.43 for each; **Figure 3**) than with one another, supporting discriminant validity between financial well-being and global life satisfaction.

### Construct and Predictive Validity

The FWB-5 Index also demonstrated construct and predictive validity (**Table 2**). For construct validity, greater childhood financial capital, participant education, and parental education were associated with higher baseline standardized FWB-5 scores, whereas greater childhood adversity was associated with lower scores. For predictive validity, a 1-SD increase in FWB-5 score was associated with fewer reported health conditions and depressive symptoms at follow-up.

## Discussion

In this study, we developed and validated a parsimonious financial well-being index derived from five indicators consistently assessed in the HRS since 2006. The FWB-5 Index demonstrated convergent validity with the CFPB-FWB Scale, discriminant validity with the Satisfaction with Life Scale, construct validity through associations with theoretically related socioeconomic and psychosocial measures, and predictive validity for subsequent health outcomes. The FWB-5 Index is intended to support longitudinal research on financial well-being, health, and aging, particularly in waves in which the CFPB-FWB Scale was not administered or is incomplete. Furthermore, this study builds on prior research on financial well-being in the HRS and further draws on established approaches to the development and validation of multidimensional measures from both the childhood socioeconomic circumstances^15^ and financial well-being psychometric^14^ literature. Specifically, we applied theoretically informed domain specification, confirmatory factor analysis, and assessments of construct and predictive validity to develop an index using existing HRS indicators.

The FWB-5 Index builds on prior work by Kezios et al.^4^ by using fewer indicators while demonstrating strong convergence with the CFPB-FWB Scale. Their eight-item index, along with a sensitivity analysis restricted to four indicators, provided support for the feasibility of a more parsimonious approach. Their item-level convergence analyses also informed our validation strategy. Notably, the FWB-5 Index exhibited a higher observed absolute Spearman correlation with the overall CFPB-FWB Scale (0.80 versus 0.65) and exceeded an absolute correlation threshold of 0.50 for more individual CFPB-FWB items (8 of 10 versus 3 of 10). Although differences in study populations and survey waves limit direct comparisons of relative performance, these findings suggest that a more parsimonious index that utilizes a theoretically motivated measurement framework can demonstrate substantial convergence with both the overall CFPB-FWB Scale and several of its individual items. That said, both indices demonstrated their weakest convergence with the items “Just getting by financially” and “Finances control my life,” suggesting that certain subjective dimensions of financial well-being may be less readily captured by existing HRS indicators or measurement approaches. Nevertheless, both approaches demonstrated convergence with the broader FWB construct, supporting the utility of existing HRS financial measures for longitudinal research on financial well-being.

Several considerations are important for the interpretation and utilization of the FWB-5 Index. As with the CFPB-FWB Scale, individual scores are most informative when interpreted comparatively rather than in isolation.^2,5,33^ Higher FWB-5 Index scores indicate greater FWB, but small differences in scores should not necessarily be interpreted as meaningful differences in lived financial circumstances. Rather, the index may be particularly useful for examining relative differences between individuals, associations with objective indicators of financial circumstances, and changes in FWB over time. This interpretation is consistent with guidance for the CFPB-FWB Scale, for which ranges of scores correspond to substantively different financial circumstances.^5,33^ For example, individuals with CFPB scores of 38–49 are classified as having “medium-low” FWB and tend to report limited savings, greater difficulty making ends meet, and greater credit constraints.^33^ In contrast, scores of 50–57 are classified as “medium-high” FWB and are associated with more favorable financial behaviors and circumstances, including greater use of automated savings and lower prevalence of financial hardship.^33^ Thus, meaningful differences in FWB are better reflected by larger shifts in the distribution than by small numerical changes.

Accordingly, in this study we standardized the FWB-5 scores to have a mean of 0 and SD of 1 and use 1-SD differences in our analyses to represent substantively meaningful variation in FWB. Depending on the research question, investigators could alternatively categorize the index using theoretically or empirically meaningful thresholds, such as below-versus above-average FWB or quantiles of the score distribution. Regardless of the approach, interpretation should emphasize meaningful differences in relative FWB rather than minor changes in the index. The FWB-5 Index may therefore be particularly useful for comparing individuals within the HRS and for examining longitudinal changes in financial well-being.

### Limitations

Several important limitations should be noted. First, we used complete-case data for both score construction and validation. Financial questions may be perceived as sensitive, stigmatizing, or uncomfortable, and nonresponse may therefore differ across population subgroups. For example, a study conducted in a primary health care setting found that, although most (73%) participants reported their income, women, racial and ethnic minority groups, caregivers of young children, and older adults were less likely to do so.^34^ Similarly, an analysis of statewide population-based postpartum survey data from the California Maternal and Infant Health Assessment (MIHA) found that respondents with missing income information tended to be younger, less educated, and of lower parity.^35^ These findings suggest that item nonresponse to financial questions may not occur randomly. Consequently, some subgroups may be underrepresented among participants with complete financial data or may provide less accurate responses, potentially introducing selection or measurement bias into the construction and validation of the FWB-5 score. In addition, our analytic sample was older and predominantly NH White, which may further limit the generalizability of our validity assessments to socioeconomically disadvantaged populations and racial and ethnic minority groups. Nevertheless, the CFPB-FWB scale itself does not incorporate demographic characteristics other than age into score calculation and recommends that scores not be calculated for individuals who do not respond to all required items.^2,5^

Second, it remains unclear whether the specific items and scoring approach used to construct the FWB-5 Index within the HRS are directly generalizable to other studies or populations. However, we developed the index using a theoretically motivated measurement framework that can be replicated and, where appropriate, recalibrated in other datasets. Additional validation in more socioeconomically and racially and ethnically diverse populations, as well as in studies with different patterns of financial-item nonresponse, is warranted.

Finally, the FWB-5 Index equally weighted the objective and subjective factor scores, assuming that both domains contribute equally to overall financial well-being. Although this approach was informed by prior work on the Childhood Financial Capital Index,^15^ this precedent does not establish that equal domain weighting is optimal for the FWB-5 Index. While our validation analyses supported the current index, they do not establish whether equal weighting is preferable to alternative approaches. Alternative weighting schemes, including those assigning greater weight to the subjective domain, warrant further investigation.

### Conclusion

In conclusion, the FWB-5 Index provides a validated, parsimonious measure of financial well-being for longitudinal research in the HRS. Like prior HRS-based indices, it offers a practical alternative when the 10-item CFPB-FWB Scale is unavailable. The index facilitates investigation of financial well-being trajectories and their associations with health and aging over extended follow-up in a longstanding cohort of older Americans.

## Data Availability

All data produced in the present study are available upon reasonable request to the authors

## Data availability

Health and Retirement Study (HRS) data may be obtained at: https://hrs.isr.umich.edu/. Details on the particular HRS datasets utilized are outlined in the methods section.

## Authors’ contributions

MH contributed to the problem definition, conducted the data analysis, and wrote the first draft. RAN obtained funding and led the problem definition, conceptualized, and supervised the implementation of the analysis plan, contributed to interpretation of the results, and reviewed and revised the manuscript. ERM, CLN, HZ, LB, OAA, KLK, RAN provided supervision of the proposal, and critically reviewed the manuscript. All authors provided critical input and insights into the development and writing of the article and approved the final manuscript as submitted.

## Declarations

Generative artificial intelligence tools were used to improve the readability of the manuscript, and all generated content was reviewed and verified by the first author (MH).

