## Supplemental Tables and Figures for "Development and Validation of a Parsimonious Financial Well-Being Index in the Health and Retirement Study"

**Supplemental Materials**

**Table S1.** Confirmatory Factor Analysis Variable Loadings and Model Fit in the FWB-5 Index CFA Sample, Health and Retirement Study, 2006–2022

**Table S2.** Participant Characteristics and Responses to Component Measures in the Convergent and Discriminant Validity Sample, Health and Retirement Study, 2020–2022.

**Table S3.** Participant Characteristics and Theoretical Correlates in the Construct and Predictive Validity Sample, Health and Retirement Study, 2006–2022.

**Figure S1.** Correlated Two-Factor Confirmatory Factor Analysis Model of the FWB-5 Index in the Validation Sample, Health and Retirement Study, 2006–2022.

**Figure S2.** Steps Used to Construct and Operationalize the FWB-5 Index.

**Table S1.** Confirmatory Factor Analysis Variable Loadings and Model Fit in the FWB-5 Index CFA Sample, Health and Retirement Study, 2006–2022.

| **Variable Factor Loadings** | **Correlated two-factor model: Train data (50% sample)** | **Correlated two-factor model: Test data (50% sample)** | **Correlated two-factor model: Full data** | **One-factor model: Train data (50% sample)** | **One-factor model: Test data (50% sample)** | **One-factor model: Full data** |
|---|---|---|---|---|---|---|
| Total Household Wealth | 0.81 | 0.79 | 0.80 | 0.43 | 0.41 | 0.42 |
| Household Income-Poverty Ratio | 0.65 | 0.65 | 0.65 | 0.36 | 0.36 | 0.36 |
| End-of-Month Financial Strain (Reverse Coded) | 0.79 | 0.81 | 0.80 | 0.79 | 0.81 | 0.80 |
| Financial Mastery | 0.53 | 0.54 | 0.53 | 0.52 | 0.53 | 0.53 |
| Satisfaction with Present Financial Situation | 0.84 | 0.81 | 0.82 | 0.82 | 0.80 | 0.81 |
| **Model Fit Statistics** | | | | | | |
| Robust Comparative Fit Index (CFI) | 0.99 | 0.99 | 0.99 | 0.82 | 0.82 | 0.82 |
| Robust Root Mean Square Error of Approximation (RMSEA) | 0.05 | 0.05 | 0.05 | 0.22 | 0.21 | 0.22 |
| Number of Observations in the Model | 11481 | 11525 | 23006 | 11481 | 11525 | 23006 |

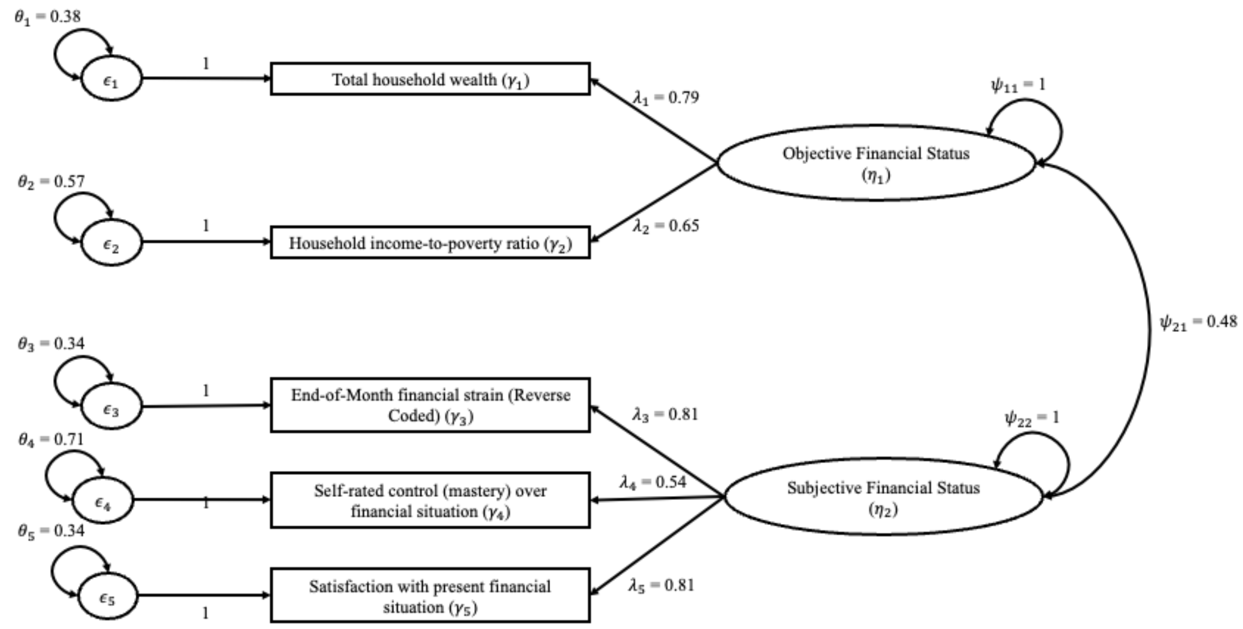


**Figure S1.** Correlated Two-Factor Confirmatory Factor Analysis Model of the FWB-5 Index in the Validation Sample, Health and Retirement Study, 2006–2022. Rectangles represent the five observed indicators ($\gamma_1$– $\gamma_5$); ovals represent the latent factors Objective Financial Status ($\eta_1$) and Subjective Financial Status ($\eta_2$); and circles labeled $\epsilon_1$– $\epsilon_5$ represent item residuals. Single-headed arrows from the latent factors to the observed indicators represent factor loadings ($\lambda_1$– $\lambda_5$), and single-headed arrows from the residual terms to the observed indicators represent residual effects. Double-headed arrows represent estimated variances ($\theta_1 - \theta_5$) and covariances ($\psi_1 - \psi_2$), including the covariance between the two latent factors ($\psi_{21}$). All parameter estimates are standardized.

**Figure S2.** Steps Used to Construct and Operationalize the FWB-5 Index.

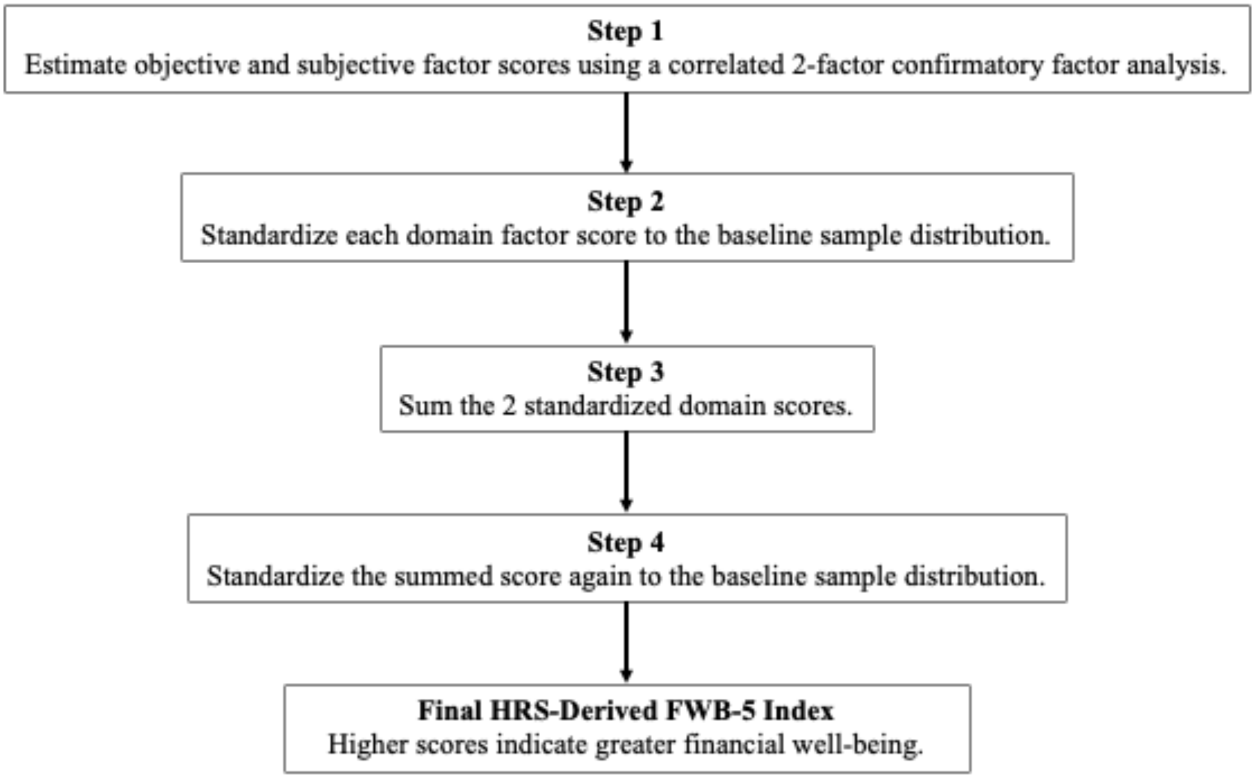

**Table S2.** Participant Characteristics and Responses to Component Measures in the Convergent and Discriminant Validity Sample, Health and Retirement Study, 2020–2022.

| **Characteristic** | **N = 7,728**[1] |
|---|---|
| HRS Year | |
| 2020 | 3,991 (52%) |
| 2022 | 3,737 (48%) |
| Age (Years) | 69 (10) |
| Male | 3,175 (41%) |
| Race/Ethnicity | |
| Hispanic | 1,049 (14%) |
| NH Black | 1,426 (18%) |
| NH White | 4,915 (64%) |
| Other | 326 (4.2%) |
| Unknown | 12 |
| Diener Satisfaction with Life Scale | 5.07 (1.47) |
| **FWB-5 Index and Subcomponents** | |
| Std. FWB-5 Index | 0.00 (1.00) |
| Std. Subjective Financial Status Factor Score | 0.00 (1.00) |
| Financial Mastery Index | 7.81 (2.32) |
| End-of-Month Financial Strain | |
| Not at all difficult | 3,932 (51%) |
| Not very difficult | 2,105 (27%) |
| Somewhat difficult | 1,310 (17%) |
| Very difficult | 287 (3.7%) |
| Completely difficult | 94 (1.2%) |
| Satisfaction with Present Financial Situation | |
| Not at all satisfied | 402 (5.2%) |
| Not very satisfied | 860 (11%) |
| Somewhat satisfied | 2,401 (31%) |
| Very satisfied | 2,468 (32%) |
| Completely satisfied | 1,597 (21%) |
| Std. Objective Financial Status Factor Score | 0.00 (1.00) |
| Total Household Wealth (2022 U.S. Dollars) | 709,259 (1,120,323) |
| Household Income-to-Poverty Ratio | 5.0 (5.0) |
| **CFPB-FWB Scale and Subcomponents** | |
| Std. CFPB-FWB Scale | 0.00 (1.00) |
| I Could Handle A Major Unexpected Expense | |
| Completely | 1,241 (16%) |
| Very Well | 1,960 (25%) |
| Somewhat | 2,571 (33%) |
| Very Little | 955 (12%) |
| Not At All | 1,001 (13%) |

| | |
|---|---|
| I Am Securing My Financial Future | |
| Completely | 1,062 (14%) |
| Very Well | 2,109 (27%) |
| Somewhat | 2,779 (36%) |
| Very Little | 1,034 (13%) |
| Not At All | 744 (9.6%) |
| Because Of My Money Situation, I Feel Like I Will Never Have The Things I Want In Life | |
| Completely | 312 (4.0%) |
| Very Well | 313 (4.1%) |
| Somewhat | 2,024 (26%) |
| Very Little | 2,397 (31%) |
| Not At All | 2,682 (35%) |
| I Can Enjoy Life Because Of The Way I Am Managing My Money | |
| Completely | 1,243 (16%) |
| Very Well | 2,368 (31%) |
| Somewhat | 2,897 (37%) |
| Very Little | 816 (11%) |
| Not At All | 404 (5.2%) |
| I Am Just Getting By Financially | |
| Completely | 637 (8.2%) |
| Very Well | 1,055 (14%) |
| Somewhat | 2,368 (31%) |
| Very Little | 1,225 (16%) |
| Not At All | 2,443 (32%) |
| I Am Concerned That The Money I Have Or Will Save Will Not Last | |
| Completely | 670 (8.7%) |
| Very Well | 499 (6.5%) |
| Somewhat | 2,662 (34%) |
| Very Little | 2,003 (26%) |
| Not At All | 1,894 (25%) |
| Giving A Gift For A Wedding, Birthday, Or Other Occasion Would Strain My Finances For The Month | |
| Always | 312 (4.0%) |
| Often | 303 (3.9%) |
| Sometimes | 1,424 (18%) |
| Rarely | 2,563 (33%) |
| Never | 3,126 (40%) |
| I Have Money Left Over At The End Of The Month | |
| Always | 2,548 (33%) |
| Often | 1,700 (22%) |
| Sometimes | 1,991 (26%) |
| Rarely | 951 (12%) |

| | |
|---|---|
| Never | 538 (7.0%) |
| I Am Behind With My Finances | |
| Always | 192 (2.5%) |
| Often | 239 (3.1%) |
| Sometimes | 898 (12%) |
| Rarely | 1,942 (25%) |
| Never | 4,457 (58%) |
| My Finances Control My Life | |
| Always | 381 (4.9%) |
| Often | 430 (5.6%) |
| Sometimes | 1,569 (20%) |
| Rarely | 2,136 (28%) |
| Never | 3,212 (42%) |

[1] n (%); Mean (SD)

**Table S3.** Participant Characteristics and Theoretical Correlates in the Construct and Predictive Validity Sample, Health and Retirement Study, 2006–2022.

| **Characteristic** | **N = 11,314**[1] |
|---|---|
| Age (Years) | 68 (10) |
| Male | 4,548 (40%) |
| Race/Ethnicity | |
| Hispanic | 886 (7.8%) |
| NH Black | 1,218 (11%) |
| NH White | 8,971 (79%) |
| Other | 238 (2.1%) |
| Unknown | 1 |
| Std. FWB-5 Index | 0.20 (1.01) |
| Childhood Financial Capital Index Z-Score | 0.05 (0.99) |
| Number of Childhood Adversities | 0.38 (0.66) |
| Participant Years of Education | 13.03 (2.96) |
| Mother's Education | 9.9 (3.5) |
| Father's Education | 9.5 (3.8) |
| Follow-up Number of Conditions Ever Reported | 2.14 (1.43) |
| Follow-up Depression (CES-D Score) | 1.26 (1.84) |

[1] Mean (SD); n (%)
